# Vitamin D recommendations in nutritional guidelines: Protocol for a systematic review, quality evaluation using AGREE-2 and analysis of potential predictors

**DOI:** 10.1101/2020.04.06.20055962

**Authors:** D Fraile-Navarro, A López-García-Franco, E Niño de Guzmán, H Pardo-Hernandez, C Canelo-Aybar, J Kuindersma, I Gich-Saladich, P. Alonso-Coello

## Abstract

**Background:** Vitamin D has been widely promoted for bone health through supplementation and fortification of the general population. However, there is growing evidence that does not support these strategies. Our aim is to review the quality and recommendations on vitamin D nutritional and clinical practice guidelines and explore predictive factors for their direction and strength.

**Methods and analysis:** We searched PubMed, EMBASE and CINAHL databases for vitamin D guidelines for the last 10 years. We aim to perform descriptive analysis, a quality appraisal using AGREE II scores (Appraisal of Guidelines Research and Evaluation) and a bivariate analysis evaluating the association recommendations and AGREE II domains’ scores and pre-specified characteristics.

**Ethics and dissemination:** This is a systematic review protocol and therefore formal ethical approval is not required, as no primary, identifiable, personal data will be collected. Patients or the public were not involved in the design of our research. However, the findings from this review will be shared with key stakeholders, including patient groups, clinicians and guideline developers. We intend to publish our results in a suitable, peer-reviewed journal.

## Introduction

Vitamin D plays a vital role in several physiological processes(1–4). The best understood of these functions is calcium regulation, counter-regulating parathyroid hormone secretion to maintain calcium serum levels(5) and calcium absorption in the gut. It is predominantly synthesized via direct sunlight (ultraviolet B radiation) exposure(6), as it is limited to only a few natural sources(7) in the human diet. Severe deficiency leads to rickets in children and osteomalacia in adults; its deficit has been associated with low bone mineral density(8), and increased risk of fractures(9).

Public health concerns about suboptimal vitamin D intake have led to the development of dietary and supplementation recommendations(10) in nutritional guidelines. However, the link between vitamin D intake and fracture risk is controversial(11). These recommendations were initially addressed to populations at risk of low sun exposure, particularly in northern latitudes(12). Early systematic reviews(13–15) showed a decrease in fractures among older institutionalized women, while more recent ones have demonstrated the absence of beneficial effect(16–22). Of note is the absence of randomized clinical trials in the general population to evaluate outcomes such as fractures, with only surrogate outcomes (e.g. bone mineral density and 25-hydroxycholecalciferol (25-OH-D)(23) being available. Vitamin D supplementation has not shown any effect in preventing cardiovascular disease or cancer(24–26), and high doses paradoxically, have been associated with an increase in falls in the elderly(27).

Another subject of debate is what constitutes vitamin D deficiency. While some studies declared epidemic proportions of deficiency(28), others have not confirmed these claims(10). Typically assessed with 25-OH-D serum levels, the threshold to maintain adequate bone health is also disputed(10).

Our aim is to review and analyze Vitamin D recommendations present in nutritional and clinical practice guidelines (CPGs) as well as potentially related factors for these recommendations.

## Methods and analysis

### Information sources & Search

We searched Medline via PubMed, EMBASE and CINAHL databases. We conducted our search for CPGs published between for the last ten years. The full search strategy can be found in **Supplementary File 1**.

### Eligibility criteria

We will include guideline publications, following the definition of the Institute of Medicine(29). We will include those that formulate recommendations for vitamin D intake, and/or screening for the general, healthy population. We will exclude guidelines only addressed to the pediatric population (to prevent rickets), guidelines on specific conditions (e.g. chronic corticoid users), guidelines addressed to osteoporotic patients exclusively, or for secondary prevention of osteoporotic fractures.

### Data extraction

We will collect: year of publication, institution, region, target population, proposed vitamin D levels, calcium advice (if given), sun exposure advice, screening advice, fortification advice, suggested methods for vitamin D obtention, and supplementation recommendations. Four reviewers will conduct screening, data extraction, and quality assessment. We will resolve disagreements through discussion or with the help of another reviewer (PA). We will classify recommendations following the distinction that the Grading of Recommendations Assessment, Development and Evaluation(GRADE) provides(30) in three categories: “Recommends”, for unequivocal, strong recommendations; “Suggests”, for weak or conditional recommendations; and “Does not recommend”, for guidelines which do not provide advice in favor or recommend against supplementation.

We will also collect information concerning the authors’ potential conflicts of interest (COIs) and how these are managed. We will focus on evaluating individual COIs reporting and the potential Institutional COIs in accordance with methodology implemented in previous evaluations(31,32). From each guideline, we will collect information on each one of the categories identified by Hakoum et al. on its methodological survey (32). These categories include not only financial conflicts of interest but other less typically acknowledged categories such as intellectual COIs and institutional financial and advocatory ties. Therefore, we will collect six different dimensions of COIs including whether there is a COI clear process reporting, if the authors reported affiliation, financial and intellectual COIs. We will also collect the type of organization that formulates the recommendations and, if available, the institution’s potential financial or advocatory COIs(32). Two reviewers (DF and CC) will collect this data independently. In case of disagreement, we will resolve it by discussion and the help of a third reviewer (PA). To perform our analysis we will summarize the different potential COIs in three categories. “Detected” for guidelines that we detected potential COIs in different of the aforementioned categories, “Undetected” for guidelines for which we do not have detected potential COIs and “Not reported” for guidelines who do not state at all a COI disclosure.

### Quality appraisal and risk of bias

We will use the AGREE II (Appraisal of Guidelines Research and Evaluation version 2)(33) instrument. This tool evaluates the methodological rigour and transparency with which a guideline is developed. After calibration, four researchers will evaluate publications independently. We will calculate the percentage of the maximum possible score for each domain, and its standardized range (from 0% to 100%).

### Data analysis

We will perform a descriptive analysis for each included guideline. We will assess the reproducibility of quantitative measurements made by different observers by calculating the intra-class correlation coefficients (ICC), and the associated 95% confidence intervals (CI) for each AGREE II domain. We will conduct a bivariate analysis to evaluate the potential association between recommendations and AGREE II scores with the following factors: region, type of organization, target population, suggested method to obtain Vitamin D, advice on sun exposure, advice on fortifying strategies, and conflicts of interest. We will include in our logistic regression model the results that remain significant in the bivariate analysis.

### Patients and Public Involvement

Patients or the public were not involved in the design of our research. However, the findings from this review will be shared with key stakeholders, including patient groups, clinicians and guideline developers.

## Data Availability

All data generated or analyzed during this study will be included in the published article (and its supplementary information files).

## List of abbreviations

AGREE II: Appraisal of Guidelines Research and Evaluation / Appraisal of Guidelines Research and Evaluation version 2
25-OH-D: 25-hydroxycholecalciferol
CPGs: Clinical Practice Guidelines
G-I-N: Guidelines International Network
GRADE: Grading of Recommendations Assessment, Development and Evaluation
COI: Conflicts of interest
ICC: Intra-class correlation coefficient
CI: Confidence interval

## Declarations

### Ethics approval and consent to participate

Not applicable.

### Authors’ contributions

DF contributed to design and developed search strategies and piloted data extraction forms and wrote this protocol. AL and PA collaborated in the design and protocol redaction. EN, EP and CC collaborated in this protocol redaction. IG collaborated in designing data analysis and redacting protocol.

